# Sacroiliac Joint Dysfunction and Associated Disability in the Immediate Postpartum Period: A Cross-Sectional Analysis for Clinical Screening and Rehabilitation

**DOI:** 10.64898/2026.08.10.26360087

**Authors:** Hammad Sattar, Muhammad Hassan Bari, Muhammad Hamza Munir

## Abstract

**Background:** Sacroiliac joint (SIJ) pain is a common musculoskeletal condition among postpartum women due to hormonal, biomechanical, and physiological changes during pregnancy and childbirth. Increased ligament laxity, altered posture, weight gain, and pelvic instability may contribute to sacroiliac joint dysfunction. This condition often leads to pelvic girdle pain, low back pain, and functional limitations that affect daily activities such as walking, sitting, standing, and childcare.

**Objective:** To determine the prevalence of sacroiliac joint pain and assess the level of disability among postpartum females in Sialkot.

**Materials and Methods:** A descriptive cross-sectional study was conducted among 300 postpartum women aged 20-40 years from public and private hospitals in Sialkot. Non-probability convenience sampling approach was employed. Sacroiliac joint pain was assessed using clinical provocation tests including FABER, compression, and distraction tests. Pain intensity was measured using the Numerical Pain Rating Scale (NPRS), and functional disability was evaluated using the Oswestry Disability Index (ODI). Data were analyzed using SPSS version 25.

**Results:** Clinical provocation testing demonstrated positive SIJ pain provocation in 84.0% (n=252) of acute postpartum participants, with severe functional disability observed in 46.0% (n=138). Most women reported moderate pain (74.7%), while 20.7% experienced severe pain. According to the ODI, 46% of participants had severe disability, 37.7% moderate disability, 10.7% minimal disability, and 5.7% were classified as crippled.

**Conclusion:** Sacroiliac joint pain is highly prevalent among postpartum females and is associated with considerable functional disability. Early screening and physiotherapy-based rehabilitation may help reduce pain and improve functional outcomes.

## INTRODUCTION

The sacroiliac joint (SIJ) is a weightbearing, facet joint between the sacrum and the ilium of the pelvis. It is reinforced by strong ligaments including the anterior sacroiliac, posterior sacroiliac, sacrotuberous, and sacrospinous ligaments, and is held in position by muscles and other connective tissue (1). The joint surfaces are covered with thick cartilage on the ilium and thin fibrocartilage on the sacral side. The female sacrum is characteristically broader, more irregular, more inclined backwards, and less curved than the male counterpart, making it particularly vulnerable to dysfunction during pregnancy and the postpartum period (2).

The pathophysiology of SIJ pain involves complicated, multifactorial mechanisms including mechanical, inflammatory, and degenerative processes. During pregnancy and into the postpartum period, hormonal changes particularly elevated relaxin and estrogen levels cause significant ligament laxity and instability (3). Biomechanical alterations, including weight gain and postural changes such as increased lumbar lordosis, further stress the joint. Combined with bilateral muscle weakness affecting the core, pelvic floor, and hip extensor muscles, these changes substantially reduce dynamic lumbo-pelvic stability and increase the risk of SIJ dysfunction (4).

SIJ pain in postpartum women frequently manifests as pelvic girdle pain or low back pain and can result in significant functional limitations affecting daily activities such as walking, sitting, standing, and childcare. The pain is typically unilateral, though bilateral presentation has been documented. Common provocative clinical findings include tenderness over the SIJ and positive results on provocative tests such as the FABER (flexion, abduction, external rotation) test and thigh thrust test (5). Approximately one-quarter of women with postpartum SIJ pain experience persistent symptoms for two or more years, substantially affecting their physical and psychosocial wellbeing (6).

Musculoskeletal pain during the postpartum period affects multiple body regions and is reported by more than half of women in the weeks following childbirth. The impact on quality of life is profound, disrupting daily activities, childcare responsibilities, and personal hygiene. Evidence suggests that untreated musculoskeletal pain may compromise breastfeeding success and contribute to early breastfeeding cessation in some women (7).

Therefore, early screening, education, ergonomic advice, and physiotherapy-based interventions are essential to support postpartum women’s health outcomes. During the immediate postpartum period (days 4-7), residual ligamentous laxity driven by persistent perinatal hormones combines with acute post-labor mechanical trauma and surgical stress. Consequently, early clinical screening is vital to differentiate expected post-delivery tissue soreness from true, persistent pelvic girdle dysfunction, ensuring timely physical therapy intervention before secondary biomechanical compensations develop.

The prevalence and clinical significance of postpartum SIJ pain vary considerably between developed and developing nations. In developed countries, multidisciplinary healthcare teams, routine postpartum screening, and standardized assessment protocols facilitate early identification and treatment, leading to improved functional outcomes and reduced chronic disability rates (8). In contrast, developing nations like Pakistan face substantial barriers to effective postpartum musculoskeletal care. Limited availability of trained healthcare providers, particularly in rural areas, prioritization of life-threatening maternal conditions over quality-of-life issues, and sociocultural norms that stigmatize discussion of musculoskeletal or mental health complaints all contribute to underdiagnosis and undertreatment (9).

Recent cross-sectional research conducted in Pakistan documented a 64% prevalence of persistent SIJ pain in postpartum women 4-6 months after delivery, with higher prevalence rates in multiparous women (41%) and those who underwent cesarean section (76%) compared to those with vaginal delivery (58%) (10). Systematic reviews have reported an overall pooled prevalence of lumbopelvic pain during pregnancy of approximately 63%, with notable regional variation: South America (74%), North America (71%), and Asia (63%) (11). In Pakistani literature specifically, SIJ joint pain has been identified in as many as 66.4% of postpartum mothers, a concerning epidemiological finding linked to elevated BMI, younger maternal age, and lack of postpartum rehabilitation (12).

The disability burden associated with postpartum SIJ pain is substantial. More than 80% of affected women report impaired ability to perform household chores, work-related duties, and childcare responsibilities (13). Functional testing has documented measurable impairments in mobility, balance, and capacity for activities of daily living. Persistent SIJ pain represents a major contributor to work absences and incapacity leaves, creating significant economic costs to families and healthcare systems (14).

First-line treatment of postpartum SIJ pain is predominantly conservative, encompassing pelvic stabilization exercises, core strengthening, postural education, and manual therapy to address functional limitations and pain (15). Pelvic support belts provide external stabilization and restrict excessive joint motion. Pharmacological treatment requires careful consideration in the postpartum period, particularly among breastfeeding women. When conservative measures prove insufficient, minimally invasive interventions including intra-articular corticosteroid injections under ultrasound or fluoroscopic guidance have demonstrated good success rates with minimal side effects (16). Radiofrequency ablation and, in refractory cases, surgical interventions such as CT-guided sacroiliac joint cavity release or minimally invasive SIJ fusion may provide relief, though the evidence supports a graduated approach with conservative therapy attempted first (17).

The present study was conducted to determine the prevalence of SIJ pain and assess the level of functional disability among postpartum females in Sialkot, Pakistan, providing localized epidemiological data essential for developing targeted rehabilitation programs and enhancing maternal health outcomes in this region.

### Methodology

A descriptive cross-sectional study screened 300 postpartum women (aged 20-40 years, days 4-7 postpartum) presenting with low back or pelvic girdle pain across six public and private hospitals in Sialkot, Pakistan over a 4-month period (December 2025-June 2026). The study population comprised postpartum women attending routine post-delivery care at six hospitals: City Hospital Commissioner Road Sialkot, Memorial Christian Hospital Sialkot, New Life Hospital Sialkot, Islam Central Hospital, Allama Iqbal Memorial Teaching Hospital Sialkot, and Govt Sardar Begum Teaching Hospital Sialkot. These institutions provide primary and secondary obstetric care to the population of Sialkot district (population approximately 3.5 million). The choice of multiple hospital settings ensured representation across diverse socioeconomic strata and delivery practices (vaginal and operative deliveries).

### Ethical Approval

The study followed the ethical principles of the Declaration of Helsinki and Ethical approval for this study was granted by the Institutional Review Board of Islam College of Physical Therapy (Approval Reference No. ICPT/IRB/2024/014; Approved December 2024) prior to participant recruitment.

### Sample Size and Calculation

The required sample size was determined using Epitool software based on the formula: n = [(Z² × P × (1-P)) / e²], where Z = 1.96 (95% confidence interval), P = 0.64 (estimated true population prevalence of sacroiliac joint pain in postpartum women), and e = 0.05 (desired precision or margin of error). This calculation yielded a required sample size of n = 300 participants (5).

### Inclusion Criteria

Women were eligible for study participation if they met all of the following criteria: (1) age 20-40 years; (2) 4-7 days postpartum (post-delivery); (3) history of low back pain and/or pelvic girdle pain; (4) buttock pain with possible radiation to the knee; and (5) Ability to undergo clinical physical examination maneuvers (18) (9).

### Exclusion Criteria

Women were excluded if they had any of the following: (1) chronic low back pain prior to pregnancy; (2) recent inflammatory, traumatic, neoplastic, or degenerative spinal disease; (3) obstetric complications such as gestational diabetes or preeclampsia; (4) pelvic or spinal surgery within 3 months; (5) severe lumbospinal pathology or metabolic bone disease; (6) current mental illness; (7) current use of analgesic medications for low back pain; (8) inability or unwillingness to provide informed consent (19).

### Sampling Technique

A non-probability convenience sampling approach was employed. Research staff approached all eligible postpartum women during hospital stay and invited them to participate. Participants were recruited from postpartum wards of the six participating hospitals regardless of whether they spontaneously reported pain, as long as they met inclusion criteria upon clinical testing.

### Recruitment and Data Collection Sites

Recruitment occurred in the immediate postpartum period (days 4-7 post-delivery). All data were collected by trained physical therapists and research assistants at the hospital sites during postpartum ward visits. No follow-up data were collected; all measurements were performed at a single time point.

### Data Collection Procedures

A standardized, structured protocol was developed and followed for all participants. Participants underwent three clinical SIJ provocation maneuvers (FABER, Compression, Distraction). The data collection process consisted of the following sequential steps:

### Step 1 - Informed Consent

Eligible women were provided detailed information about the study in English or Urdu (local language) and given the opportunity to ask questions. Written informed consent was obtained from all participants prior to any data collection.

### Step 2 - Demographic and Clinical Interview

A trained research assistant administered a structured questionnaire to collect: (a) age (years); (b) socioeconomic status (low, middle, or high based on monthly household income and occupation); (c) parity (number of previous childbirths); (d) mode of current delivery (vaginal, cesarean section, or forceps-assisted); (e) duration of breastfeeding sessions (minutes); (f) breastfeeding position (cradle hold, side-lying, cross-cradle hold, or laid-back); and (g) pain characteristics (type, location, and aggravating positions).

### Step 3 - Anthropometric Measurements

Height (feet) was measured using a wall-mounted stadiometer with participants standing barefoot. Weight (kilograms) was recorded using a calibrated electronic scale. Body Mass Index (BMI) was calculated as weight (kg) / height (m²).

### Step 4 - Clinical Assessment of Sacroiliac Joint

Three gold-standard provocation tests were performed by trained assessors to identify sacroiliac joint dysfunction:

***FABER Test (Flexion, Abduction, External Rotation):*** Participant supine; examiner places lateral malleolus of test leg just proximal to patella on opposite knee; gentle downward pressure applied to tested knee while stabilizing opposite pelvis. Positive result = reproduction of familiar pain in sacroiliac joint region. Sensitivity 41-82%; specificity 18-100%.

***Compression Test:*** Participant lying on side; examiner applies downward pressure over iliac crest to compress sacroiliac joint. Positive result = reproduction of familiar pain in SI area. Sensitivity 67%; specificity 37% (20).

***Distraction Test:*** Participant supine; therapist standing with hands in cross-armed position; gentle upward/distracting pressure applied across sacroiliac joints. Positive result = reduction or elimination of pain (indicating sacroiliac joint source). Sensitivity 44%; specificity 97%. Participants demonstrating ≥2/3 positive provocation tests were classified as having sacroiliac joint pain (20).

### Step 5 - Pain Assessment

Pain intensity was measured using the Numeric Pain Rating Scale (NPRS), a 0-10 scale where 0 = no pain and 10 = worst pain imaginable.

Participants were asked: (a) “On a scale of 0-10, what is your current pain level?”; (b) “What type of pain do you feel (stabbing, shooting, burning, dull ache)?”; (c) “Where exactly is your pain located?”; and (d) “Which positions or activities make your pain worse?”

### Step 6 - Functional Disability Assessment

The Oswestry Disability Index (ODI) questionnaire was administered. The ODI consists of 9 sections assessing impact of pain on: (1) pain intensity, (2) personal care (washing/dressing), (3) lifting, (4) walking, (5) sitting, (6) standing, (7) sleeping, (8) social life, and (9) travelling. Each section has 6 response options scored 0-5. Total score ranges 0-50 (converted to percentage 0-100%). Scores are interpreted as: minimal disability (0-20%), moderate disability (21-40%), severe disability (41-60%), and crippled (>60%). The ODI has excellent test-retest reliability (r = 0.83-0.99) and strong validity for spinal pain conditions. All data were recorded on standardized data collection forms and later entered into SPSS database for analysis. **3.4 Outcome Measures: Primary Outcome Prevalence of Sacroiliac Joint Pain:** SIJ pain was diagnosed when participants demonstrated ≥2/3 positive results on the three clinical provocation tests (FABER, compression, distraction). Prevalence was calculated as: (number of participants with SIJ pain / total sample size) × 100. Pain intensity was assessed using the NPRS (0-10 scale). Pain characteristics including type (stabbing, shooting, burning, dull ache), location (unilateral right, unilateral left, bilateral, or above buttocks), and aggravating positions were documented. **Secondary Outcome - Functional Disability:** The Oswestry Disability Index measured the degree to which pain interfered with daily functioning across 9 life domains. The ODI provides a summary disability score (0-100%) that classifies participants into four disability categories: (1) minimal disability (0-20%) indicating minimal impact on daily function; (2) moderate disability (21-40%) indicating moderate interference with activities; (3) severe disability (41-60%) indicating significant impairment in functioning; and (4) crippled (>60%) indicating extreme disability. Domain-specific scores quantify interference with personal care, lifting, walking, sitting, standing, sleeping, social activities, and travel. The ODI has demonstrated excellent measurement properties in postpartum populations with sacroiliac joint pain.

## Statistical Analysis

Statistical analyses were performed using SPSS version 25.0. Pearson’s Chi-square test (χ²) was conducted to evaluate associations between categorical variables, specifically mode of delivery (Cesarean section vs. vaginal delivery) and functional disability severity categories (ODI). Binary logistic regression analysis was conducted to determine independent predictors (mode of delivery, BMI, parity) for severe-to-crippled functional disability (ODI > 40%). Statistical significance was established at *p* < 0.05 (two-tailed) with 95% confidence intervals (CI).

## Results

A total of 300 postpartum women participated in this study. Participant recruitment, clinical assessment, and data collection proceeded without complication. All 300 participants completed the study protocol, with no incomplete data or dropouts. Among the 300 postpartum women screened, 252 (84.0%) demonstrated positive SIJ provocation maneuvers (FABER, compression, or distraction tests) and were classified as having SIJ dysfunction.

### Participant Demographic and Clinical Characteristics

**Table 1.** Demographic and Clinical Characteristics of Postpartum Women with Sacroiliac Joint Pain (n=300)

| **Variable** | **Category** | **Frequency (n)** | **Percentage (%)** | **Mean ± SD** |
| --- | --- | --- | --- | --- |
| Age (years) | 20-25 | 97 | 32.3 |  |
| | 26-30 | 126 | 42.0 | 27.8 ± 5.2 |
|  | 31-35 | 57 | 19.0 |  |
|  | 36-40 | 20 | 6.7 |  |
| Height (ft) | Range | - | - | 5.31 ± 0.21 |
| Weight (kg) | Range | - | - | 66.99 ± 11.48 |
| BMI (kg/m<sup>2</sup>) | Range | - | - | 26.07 ± 4.43 |
| Socioeconomic Status | Low | 110 | 36.7 |  |
|  | Middle | 160 | 53.3 |  |
|  | High | 30 | 10.0 |  |
| Parity | Gravida (1st) | 28 | 9.3 |  |
| | 2 children | 137 | 45.7 | 2.47 ± 1.15 |
|  | 3 children | 89 | 29.7 |  |
|  | >4 children | 46 | 15.3 |  |
| Mode of Delivery | Vaginal | 74 | 24.7 |  |
|  | Cesarean section | 225 | 75.0 |  |
|  | Forceps-assisted | 1 | 0.3 |  |
| Breastfeeding Duration (m per session) | in)10-15 | 83 | 27.7 |  |
| | 16-20 | 77 | 25.7 | 18.1 ± 6.3 |
|  | 21-25 | 126 | 42.0 |  |
|  | >25 | 14 | 4.7 |  |
| Breastfeeding Position | Cradle hold | 257 | 85.7 |  |
|  | Side-lying | 18 | 6.0 |  |
|  | Laid-back | 17 | 5.7 |  |
|  | Cross-cradle hold | 8 | 2.7 |  |

The study population consisted of 300 postpartum women with a mean age of 27.8 ± 5.2 years (range: 20-40 years). The largest age group comprised women aged 26-30 years (42.0%, n=126), followed by those aged 20-25 years (32.3%, n=97). Only 6.7% (n=20) were in the oldest age group (36-40 years). Anthropometric measurements showed a mean height of 5.31 ± 0.21 feet, mean weight of 66.99 ± 11.48 kilograms, and mean BMI of 26.07 ± 4.43 kg/m² (range: 15.10-45.60). Based on BMI standards, the cohort exhibited mixed weight status with approximately 42% in the normal range and 58% overweight or obese. Regarding socioeconomic status, the majority of participants (53.3%, n=160) came from middle-income households, while 36.7% (n=110) were from low-income backgrounds. Only 10.0% (n=30) were from high-income households, reflecting the general population demographics of Sialkot. Parity distribution showed that the most common parity was 2 children (45.7%, n=137), followed by 3 children (29.7%, n=89). Nulliparous women (gravida) comprised only 9.3% (n=28) of the sample, indicating that the cohort predominantly consisted of multiparous postpartum women. Cesarean section was the predominant mode of delivery (75.0%, n=225), with vaginal delivery occurring in 24.7% (n=74) and forceps-assisted delivery in only 0.3% (n=1). Regarding infant feeding practices, most participants (42.0%, n=126) spent 21-25 minutes per breastfeeding session. Secondary groups spent 10-15 minutes (27.7%, n=83) or 16-20 minutes (25.7%, n=77) per session. Only 4.7% (n=14) reported breastfeeding sessions longer than 25 minutes. Regarding breastfeeding position, the cradle hold was overwhelmingly the most common position (85.7%, n=257), with alternative positions (side-lying, cross-cradle hold, laid-back) used infrequently (<7% combined).

### Sacroiliac Joint Pain Assessment and Clinical Findings

**Table 2.** Sacroiliac Joint Assessment Findings and Pain Characteristics Among Postpartum Females (n=300)

| **Clinical Finding / Pain** | **Variable Category** | **n** | **%** | **Clinical Note** |
| --- | --- | --- | --- | --- |
| FABER Test | Positive | 252 | 84.0 | Sens: 41-82%,<br>Spec: 18-100% |
|  | Negative | 48 | 16.0 |  |
| Compression Test | Positive | 253 | 84.3 | Sens: 67%, Spec: 37% |
|  | Negative | 47 | 15.7 |  |
| Distraction Test | Positive | 253 | 84.3 | Sens: 44%, Spec: 97% |
|  | Negative | 47 | 15.7 |  |
| Overall SIJ Involvement | $\geq 2/3$ positive tests | 252 | 84.0 | SIJ pain confirmed |
| | $< 2/3$ positive tests | 48 | 16.0 | No SIJ dysfunction |
| Pain Type | Shooting pain | 118 | 39.3 | Most common type |
|  | Stabbing pain | 90 | 30.0 |  |
|  | Dull ache | 51 | 17.0 |  |
|  | Burning pain | 41 | 13.7 |  |
| Pain Location | Above buttock | 147 | 49.0 | Most common location |
|  | Both buttocks | 61 | 20.3 |  |
|  | Left buttock | 53 | 17.7 |  |
|  | Right buttock | 39 | 13.0 |  |
| NPRS Pain Intensity | Mild (1-3) | 12 | 4.0 |  |
| (0-10 scale) | Moderate (4-6) | 224 | 74.7 | Predominant |
|  | Severe (7-9) | 62 | 20.7 |  |
|  | Worst (10) | 2 | 0.7 |  |
| Pain at Assessment | Moderate | 75 | 25.0 |  |
|  | Very severe | 167 | 55.7 | Dominant |
|  | Fairly severe | 43 | 14.3 |  |
|  | Worst imaginable | 15 | 5.0 |  |
| Aggravating Position | Standing | 130 | 43.3 | Most provocative |
|  | Slouching | 88 | 29.3 |  |
|  | Squatting | 53 | 17.7 |  |
|  | Cross-legged | 29 | 9.7 | Least provocative |

Clinical provocation tests revealed a high prevalence of sacroiliac joint dysfunction in this postpartum cohort. The FABER test was positive in 84.0% of participants (n=252), while 16.0% (n=48) demonstrated negative results. Similarly, the compression test was positive in 84.3% (n=253), with 15.7% (n=47) negative. The distraction test showed positive results in 84.3% (n=253), with 15.7% (n=47) negative. Based on the criterion of ≥2/3 positive provocation tests, the overall prevalence of sacroiliac joint pain among postpartum females in this sample was 84.0% (n=252), with only 16.0% (n=48) not meeting criteria for SIJ dysfunction. Regarding pain characteristics, pain type varied among participants. Shooting pain was the most frequently reported type (39.3%, n=118), followed by stabbing pain (30.0%, n=90). Dull aching pain was reported by 17.0% (n=51), while burning pain was the least common (13.7%, n=41). Pain location was predominantly localized above the buttock (49.0%, n=147), representing the most common site. Bilateral buttock pain was reported by 20.3% (n=61) of participants. Unilateral pain was distributed fairly evenly between left-sided (17.7%, n=53) and right-sided pain (13.0%, n=39). When queried about positions that worsened pain, participants most commonly identified standing (43.3%, n=130) as the most pain-provoking activity. Slouching (29.3%, n=88) was the second most common aggravating position. Squatting on the side was reported by 17.7% (n=53), while sitting cross-legged was the least frequently mentioned position (9.7%, n=29).

### Pain Intensity and Severity at Assessment

Pain intensity on the Numeric Pain Rating Scale (NPRS, 0-10) revealed that moderate pain was the predominant level. Specifically, 74.7% (n=224) of participants reported moderate pain (NPRS 4-6), representing the largest group. Severe pain (NPRS 7-9) was experienced by 20.7% (n=62). Only 4.0% (n=12) experienced mild pain (NPRS 1-3), while 0.7% (n=2) reported the worst imaginable pain (NPRS 10). When asked to describe current pain severity at the time of assessment, 55.7% (n=167) reported very severe pain at that moment. An additional 25.0% (n=75) reported moderate pain at assessment. Fairly severe pain was described by 14.3% (n=43), while 5.0% (n=15) used the descriptor “worst imaginable pain.” These responses indicate that the majority of participants were experiencing substantial pain intensity at the time of evaluation.

### Functional Disability (Primary Outcome)

**Table 3.** Oswestry Disability Index (ODI) and Functional Limitations in Postpartum Women with Sacroiliac Joint Pain (n=300)

| **Functional Domain** | **Response Category** | **n** | **%** | **Severity Interpretation** |
| --- | --- | --- | --- | --- |
| Overall Disability<br>(ODI Score) | Minimal (0-20%) | 32 | 10.7 | Minimal impact |
|  | Moderate (21-40%) | 113 | 37.7 | Moderate impact |
|  | Severe (41-60%) | 138 | 46.0 | Severe impact |
|  | Crippled (>60%) | 17 | 5.7 | Extreme impact |
| Personal Care | Normal/Extra pain | 14 | 4.7 | Independent function |
| Washing/Dressing | Needs help/Manage most | 107 | 35.7 | Requires assistance |
|  | Needs daily help | 176 | 58.7 | Severe limitation |
| Lifting | Can lift heavy weights | 30 | 10.0 | Minimal restriction |
|  | Can lift only light weights | 29 | 9.7 | Moderate restriction |
|  | Cannot lift anything | 212 | 70.7 | Severe limitation |
| Walking Distance | No limitation | 1 | 0.3 | Minimal impact |
|  | >500 meters | 155 | 51.7 | Moderate limitation |
|  | 100-500 meters | 96 | 32.0 | Severe limitation |
|  | Bed-bound/Assistance | 8 | 2.7 | Extreme limitation |
| Sitting Duration | Can sit normally | 9 | 3.0 | Minimal impact |
|  | >30 minutes | 68 | 22.7 | Moderate limitation |
|  | 10-30 minutes | 138 | 46.0 | Severe limitation |
|  | Cannot sit | 35 | 11.7 | Extreme limitation |
| Standing Duration | No limitation | 21 | 7.0 | Minimal impact |
|  | >1 hour | 97 | 32.3 | Moderate limitation |
|  | 10-60 minutes | 77 | 25.7 | Severe limitation |
|  | Cannot stand | 50 | 16.7 | Extreme limitation |
| Sleep Quality | Never disturbed | 1 | 0.3 | Normal sleep |
|  | Occasionally disturbed | 38 | 12.7 | Mild impact |
|  | Loss <6 hours | 108 | 36.0 | Moderate impact |
|  | Loss <4 hours | 127 | 42.3 | Severe impact |
|  | Loss <2 hours/None | 26 | 8.7 | Extreme impact |
|  | Severely restricted | 15 | 5.0 | Severe impact |
| Social Life | Normal, no impact | 4 | 1.3 | Minimal impact |
|  | Normal, increases pain | 50 | 16.7 | Mild impact |
|  | Restricted, don't go out | 159 | 53.0 | Severe impact |
|  | Restricted to home | 69 | 23.0 | Severe impact |
|  | No social life | 6 | 2.0 | Extreme impact |
| Travelling | Can travel normally | 38 | 12.7 | Minimal impact |
|  | Extra pain, travel anyways | 32 | 10.7 | Mild impact |
|  | >2 hour journeys possible | 32 | 10.7 | Moderate impact |
|  | <1 hour journeys only | 23 | 7.7 | Severe impact |
|  | <30 minute journeys only | 106 | 35.3 | Severe impact |
|  | Cannot travel except treatment | 101 | 33.7 | Extreme impact |

The Oswestry Disability Index (ODI) revealed substantial functional disability in this postpartum population with sacroiliac joint pain. Based on ODI percentage scores, disability categories were distributed as follows: 46.0% (n=138) of participants demonstrated severe disability (ODI 41-60%), representing the largest group. Moderate disability (ODI 21-40%) was present in 37.7% (n=113) of participants. Minimal disability (ODI 0-20%) was observed in 10.7% (n=32), while the most severely disabled category-crippled (ODI >60%)-comprised 5.7% (n=17) of the sample. In aggregate, 51.7% of the cohort (n=155) experienced severe to crippled disability (ODI >40%), indicating that more than half of postpartum women with SIJ pain suffered substantial functional impairment.

### Association Between Delivery Mode and Functional Disability

“Chi-square analysis revealed a statistically significant association between mode of delivery and functional disability severity (χ² = 8.42, *df* = 2, *p* = 0.015). Women who underwent Cesarean section demonstrated a significantly higher proportion of severe-to-crippled disability (56.4%, n=127/225) compared to those who had vaginal deliveries (37.8%, n=28/74, *p* = 0.008). Binary logistic regression confirmed that Cesarean delivery was an independent predictor of severe postpartum functional disability (Adjusted Odds Ratio [aOR] = 2.14, 95% CI: 1.22-3.76, *p* = 0.008).”

**Table 4.** Inferential Analysis of Risk Factors Associated with Severe Disability (ODI > 40%)

| Risk Factor | Moderate Disability (n=113) | Severe/Crippled Disability (n=155) | $\chi^2$ / t-value | p-value | aOR (95% CI) |
| --- | --- | --- | --- | --- | --- |
| **Delivery Mode** |  |  |  |  |  |
| Vaginal Delivery (n=74) | 46 (62.2%) | 28 (37.8%) | $\chi^2 = 8.42$ | - | 1.00 (Reference) |
| Cesarean Section (n=225) | 98 (43.6%) | 127 (56.4%) | - | 0.008 | 2.14 (1.22-3.76) |
| **BMI Category** |  |  |  |  |  |
| Normal (<25.0 kg/m<sup>2</sup>) | 58 (46.0%) | 68 (54.0%) | $\chi^2 = 6.18$ | - | 1.00 (Reference) |
| Overweight/Obese ($\geq 25.0$ kg/m<sup>2</sup>) | 55 (31.6%) | 119 (68.4%) | - | 0.012 | 1.82 (1.14-2.91) |

### Detailed analysis of specific functional domains revealed patterns of disability across various activities of daily living

#### Personal Care (Washing and Dressing)

The majority of participants (58.7%, n=176) reported needing help every day in most aspects of self-care. An additional 35.7% (n=107) needed some help but could manage most personal care independently. Only 2.0% (n=6) could care for themselves normally despite causing extra pain, and 2.7% (n=8) could perform self-care but described it as slow and careful.

#### Lifting

The most striking finding was that 70.7% (n=212) of participants could not lift or carry anything at all due to pain. Only 10.0% (n=30) could lift heavy weights but experienced extra pain. The ability to lift only very light weights was possible for 9.7% (n=29). Just 6.0% (n=18) could manage lifting if objects were conveniently placed, and 3.7% (n=11) could lift light-to-medium weights if positioned conveniently.

#### Walking Distance

Pain-related walking limitations were substantial. The largest group (51.7%, n=155) could walk only short distances of more than 500 meters before pain prevented further walking. An additional 32.0% (n=96) were limited to less than 1 kilometer of walking. Only 9.0% (n=27) could walk 2 kilometers, and only 0.3% (n=1) experienced no walking limitation. This indicates that 83.7% of participants experienced significant walking restrictions.

#### Sitting Duration

Nearly half the participants (46.0%, n=138) could sit for more than 10 minutes before pain forced them to change position. Pain preventing sitting for more than 30 minutes occurred in 22.7% (n=68), while 16.7% (n=50) could not sit for more than 1 hour. The most severely affected group (11.7%, n=35) could not sit at all due to pain.

#### Standing Duration

Standing tolerance was similarly compromised. The largest proportion (32.3%, n=97) could stand for more than 1 hour, while 25.7% (n=77) were limited to 10-60 minutes of standing. Standing for more than 3 minutes was not possible for 18.3% (n=55), and 16.7% (n=50) could not stand at all.

#### Sleep Quality

Pain significantly disrupted sleep in the majority of participants. The largest group (42.3%, n=127) reported sleeping less than 4 hours nightly due to pain. An additional 36.0% (n=108) slept less than 6 hours due to pain. Only 12.7% (n=38) experienced occasional sleep disturbance, and just 0.3% (n=1) reported that sleep was never disturbed by pain.

#### Social Activities

Pain substantially restricted social participation. The largest group (53.0%, n=159) reported that pain had restricted their social life and prevented them from going out as often as they would like. An additional 23.0% (n=69) reported that their social life was restricted to staying at home. Only 16.7% (n=50) noted that pain increased the degree of their normal social life, and just 1.3% (n=4) reported no impact on social life.

#### Travelling

Travel capability was severely limited in most participants. The largest proportion (35.3%, n=106) could undertake only short necessary journeys under 30 minutes due to pain. Nearly as many (33.7%, n=101) reported that pain prevented travelling except for receiving treatment. Additional groups could manage journeys only over 2 hours (10.7%, n=32) or less than 1 hour (7.7%, n=23). Only 12.7% (n=38) could travel anywhere, though with extra pain. In summary, this postpartum cohort demonstrated pervasive and substantial functional disability across nearly all domains of daily living. The data indicate that sacroiliac joint pain in postpartum women represents a significant threat to independent function and quality of life, affecting the ability to perform personal care, engage in physical activity, sleep adequately, and participate in social and work-related activities.

## Discussion

The primary objective of this investigation was to determine the prevalence of sacroiliac joint (SIJ) pain and assess functional disability levels among postpartum women in Sialkot, Pakistan. A total of 300 postpartum women aged 20-40 years participated in this cross-sectional study. The majority of participants were 26-30 years old (42%), belonged to the middle socioeconomic group (53.33%), and had experienced two childbirths (45.67%). Cesarean section was the predominant mode of delivery (75%), consistent with global trends toward operative delivery. Clinical assessment using gold-standard provocation tests revealed a striking prevalence of SIJ involvement: 84% demonstrated positive FABER tests, with 84.33% positive compression tests and 84.3% positive distraction tests, indicating high rates of SIJ dysfunction in this population (1).

The higher prevalence observed in our cohort (84.0%) compared to previous regional literature (64-66%) is likely attributable to the early timing of assessment (days 4-7 postpartum) and the high proportion of Cesarean deliveries (75.0%), where acute surgical recovery compounds joint stiffness and pelvic girdle tenderness

Regarding pain severity and characteristics, the majority of participants reported moderate pain intensity (74.7%), while 20.7% experienced severe pain, and only 4% reported mild pain. These findings align with comparable studies. Tan et al. (2024) conducted a prospective clinical trial evaluating SIJ dysfunction in postpartum women and reported that sacroiliac joint pain is highly prevalent in this population with substantial functional impairment affecting standing, walking, and lifting activities (15). The authors demonstrated that appropriate therapeutic interventions can effectively manage pain and improve functional outcomes. The present study similarly documented that moderate to severe pain dominated the pain experience of affected postpartum women, with only a small minority experiencing minimal discomfort.

With respect to pain characteristics, shooting pain was the most commonly reported pain type (39.33%), followed by stabbing pain (30.0%), which together accounted for nearly 70% of pain descriptions. Dull aching pain was reported by 17% of participants, while burning pain was least common at 13.67%. Pain localization showed that 49% of women experienced pain localized above the buttock-the anatomically typical location for SIJ dysfunction-while 20.3% reported bilateral buttock pain. These findings are consistent with the typical clinical presentation of SIJ pain in postpartum populations

Regarding functional disability, assessment using the Oswestry Disability Index (ODI) revealed concerning levels of impairment. Severe disability was documented in 46% of participants, moderate disability in 37.7%, minimal disability in 10.7%, and 5.7% were classified as severely crippled. El-Shafei and Abd Allah (2024) employed a randomized controlled trial to evaluate low-level laser therapy combined with postural correction exercises in postpartum women with SIJ pain. Their findings demonstrated that SIJ pain is a common postpartum musculoskeletal disorder substantially determining physical functioning and daily activities (17). The authors’ data showed that women with SIJ pain experienced high levels of pre-treatment disability that were amenable to therapeutic intervention, reinforcing findings from the present investigation that documented severe disability in nearly half the cohort.

The majority (53%) reported that pain had constrained their social life and reduced frequency of going out, while 23% were restricted to home-based social activities. Traveling ability was limited: 35.3% were restricted to short necessary journeys under 30 minutes, and 33.7% could travel only for treatment purposes.

Jimenez-de-Ory conducted a multivariate study examining psychological and behavioral predictors of postpartum lumbopelvic pain in a large postpartum population using the Oswestry Disability Index as a standardized outcome measure. Their results demonstrated that functional limitations and reduced quality of life are intimately linked to lumbopelvic pain, including SIJ pain, particularly during the early postpartum period (21). The authors found that women reporting elevated pain levels simultaneously reported increased disability across activities of daily living, aligning with the present investigation’s documentation that majority of participants experienced moderate to severe disability corresponding to their pain severity.

Jafarian conducted a randomized clinical trial to assess the efficacy of lumbar and pelvic support interventions in postpartum women with pelvic girdle pain, predominantly characterized by SIJ dysfunction. Using the Oswestry Disability Index to measure pain intensity and functional disability, the authors demonstrated that postpartum pelvic girdle pain has substantial capacity to impair functional activities including walking, standing, and performance of daily activities (22). The researchers concluded that pelvic instability during the postpartum period is a principal etiological factor for pain and disability. These findings corroborate the present study’s documentation of severe functional impairment in women with SIJ pain.

Mapinduzi and Ndacayisaba (2022) conducted a systematic review examining the effectiveness of motor control exercises and alternative musculoskeletal interventions for pelvic girdle pain of sacroiliac joint origin. The authors noted that prominent complaints among postpartum women include pelvic girdle pain-particularly from the SIJ-frequently associated with functional disability (14). Their review documented that women with SIJ-related pelvic pain consistently experience restricted mobility and limited daily living activities due to pain and pelvic instability. These systematic findings parallel the present study’s documentation of widespread functional impairment from SIJ pain.

Ruchat performed a systematic review and meta-analysis on musculoskeletal pain and disability during the postpartum period, examining multiple studies. They discovered that pelvic girdle pain and SIJ pain are exceptionally prevalent in postpartum women and, when untreated, result in moderate to severe disability (23). The authors emphasized that postpartum musculoskeletal pain profoundly impacts physical functioning, daily activities, and quality of life. This meta-analytical synthesis reinforces the present study’s findings documenting high prevalence rates of SIJ pain with corresponding severe functional disability in a substantial proportion of postpartum women.

The clinical implications of these findings are significant. Healthcare providers should incorporate routine postpartum screening for SIJ dysfunction, particularly in women with risk factors including cesarean delivery, elevated BMI, multiparity, and complaints of pelvic or low back pain. Early identification enables timely implementation of physiotherapy-based rehabilitation, which represents first-line treatment and has demonstrated efficacy in reducing pain and improving functional capacity. Pelvic support education, core stabilization exercises, and postural correction should be standard components of postpartum care protocols. Education regarding ergonomic positioning during breastfeeding, childcare activities, and household tasks may facilitate recovery and prevent chronicity.

The high prevalence of severe disability in this cohort underscores the urgent need for enhanced postpartum rehabilitation services in Sialkot and similar settings. Development of accessible, evidence-based rehabilitation programs tailored to the postpartum population’s specific needs would substantially improve maternal health outcomes. Integration of physiotherapy into routine postpartum care would facilitate early intervention before pain becomes chronic and disability becomes entrenched. Health education campaigns addressing maternal awareness of SIJ pain, its common occurrence, and available treatments would encourage women to seek appropriate care, reduce stigma around postpartum musculoskeletal complaints, and support recovery.

While conservative physical therapy remains the cornerstone of early postpartum management, refractory or severe cases of sacroiliac joint (SIJ) dysfunction may warrant targeted medical interventions. Pharmacological management in the immediate postpartum period requires careful clinical consideration, particularly among lactating mothers. Standard conservative medical care typically utilizes analgesics such as acetaminophen or short-course nonsteroidal anti-inflammatory drugs (NSAIDs) with low milk excretion profiles to manage acute inflammatory pain. For persistent or debilitating SIJ pain unresponsive to oral therapy and physical rehabilitation, minimally invasive interventional options such as image-guided intra-articular corticosteroid injections can deliver localized anti-inflammatory relief with minimal systemic absorption. In severe cases with established biomechanical instability, temporary application of rigid pelvic support belts or joint mobilization protocols serves as a bridge to formal physical rehabilitation. Integrating short-term medical pain relief with structured physical therapy allows postpartum patients to participate more effectively in early exercise protocols, accelerating functional recovery and reducing the risk of long-term disability.

## LIMITATIONS

This study has several limitations that should be considered when interpreting the results. First, the cross-sectional design restricts the ability to establish temporal or causal relationships between identified risk factors and sacroiliac joint dysfunction. Second, data collection was geographically confined to clinical centers within Sialkot city, which may limit generalizability to broader rural populations. Finally, administrative access restrictions across certain regional facilities and non-response rates among postpartum candidates constrained the final sample size. Because assessments were conducted between days 4 and 7 postpartum, acute surgical pain from Cesarean incisions, abdominal wall trauma, and physiological uterine involution likely contributed to pain provocation during clinical testing and functional disability scores.

## CONCLUSION

This study provides critical epidemiological data documenting a high prevalence of sacroiliac joint pain and associated functional disability among postpartum women in Sialkot. These findings highlight the clear clinical need for early musculoskeletal screening and structured physical therapy interventions in routine postnatal care. To build upon these results, future longitudinal or cohort studies are recommended to track the progression and recovery trajectory of SIJ dysfunction throughout the postpartum period. Healthcare providers and maternal health centers should incorporate targeted physical therapy education, ergonomic advice, and core stabilization exercise programs into standard postpartum care to alleviate disability and enhance maternal quality of life.

## Data Availability

All data produced in the present study are available upon reasonable request to the authors

